# A classification model to predict specialty drug use

**DOI:** 10.1101/2021.06.30.21259718

**Authors:** Xianglian Ni, Andrew Fairless, Jasmine M. McCammon, Farbod Rahmanian, Heather Lavoie

## Abstract

**Objective:** Predicting who is likely to become utilizers of specialty drugs allows care managers to have an early intervention and payers to have financial preparation for the upcoming spending. Our administrative claims-based predictive model is to predict the members who might use specialty drugs.

**Materials and Methods:** A national database* and a commercial health plan claim data were used to select a total 6.5 million people who were not taking any specialty drugs before the Target Prediction Window. There were about 136,700 members who were older than 65 in the study. We extracted 81 features from past history of medical, pharmacy claims, and demographic data to predict the specialty drug use in the following year. Members having at least three-month continuous enrollment either under medical or pharmacy plan in the previous year immediately before the start of the target prediction window and with no specialty drug taking history were eligible for this study. We trained and tuned on 75% of the data using an extreme gradient boosting binary classifier. We used the remaining 25% of the data to predict the outcomes and evaluate the performance. We also recorded the performance for the age group older than 65 years old.

**Results:** There were 3% of members who used specialty drugs in the cohort under the current study. The important features for prediction included age, monthly pharmacy payment, monthly medical payment, diseases, procedure, or drug-related codes. On the test data with members of all ages, model performance for the area under the receiving operator characteristics curve (AUROC) was 78.6%. For the test set on members older than 65 (prevalence rate 3.6%), we had an AUROC of 79.3%.

**Discussion:** There is no similar machine learning model in the field to predict specialty drug use. Our model provides an unparalleled opportunity to allow early intervention for people who might develop diseases that require specialty drug use. It is also important for health plans and providers to know their covered population who might use specialty drugs and predict the increased cost in the next year.

**Conclusion:** A predictive model of specialty drug use can be helpful for both payers and providers to prepare for a spending spike or have an early intervention. In return, this helps to improve patients’ overall satisfaction.

## BACKGROUND AND SIGNIFICANCE

Specialty drugs are considered high-cost, high-complexity, or high-touch medications. Patients who use specialty drugs usually have complex chronic diseases and life-altering conditions. Many innovative specialty drugs are entering the health care market at a rapid rate. Some specialty drugs are the only effective medications for the treatment of certain diseases or conditions. Given the complexity of these drugs, patients taking specialty drugs often rely on active clinical service and management to have medication access, ensure safe use, and achieve optimal therapeutic outcomes [1].

In addition to their unique status of treating certain complex chronic diseases, specialty drugs have received increased attention by payers, providers, patients, and even policymakers due to their high prices and aggregate impact on total health care costs [2]. In retail pharmacies, specialty drugs accounted for only a small portion of total prescriptions dispensed. However, they made up to 37.7% of retail and mail-order prescription spending net of rebates in 2016-2017 [3]. About 2% of the population used specialty drugs in 2019, yet their share of spending accounted for 47.7% of total prescription drug expenditures [4].

Specialty drugs are one of the fastest growing cost areas in the health care sector. Although small molecules continue to play an important role in innovative treatments, the pharmaceutical industry is shifting its focus from small molecules to specialty medicines. In the following decade, the high cost of new specialty drugs will significantly impact patients and health plans. For some chronic conditions, a year of treatment with a specialty drug can cost a patient more than $100,000 [2]. Notably, patients treated with specialty drugs have to face high out-of-pocket (OOP) costs. Health plans usually require coinsurance payment, and that payment can be as high as 33% [5]. Increased drug cost also significantly impacts payers as they need to maintain a balance between costs and revenue. The high cost of drugs has put an increasing burden on payers’ budgets [6].

Specialty drugs can be covered under either pharmacy or medical plans. In 2019, approximately 40% of specialty drug costs were paid as a medical benefit, and 60% were paid as a pharmacy benefit. The top three therapeutic classes of total specialty spend under medical and pharmacy claims combined included autoimmune disease, oncology, and multiple sclerosis. Most specialty drugs for autoimmune diseases were dispensed under the pharmacy benefit, and those drugs comprise 32% of total specialty spend. The oncology specialty drugs were mostly covered under the medical benefit and made up to 25% of total specialty costs [7].

For providers, the ability to predict who might use specialty drugs among their covered population allows them to implement an early intervention, which can promote patient participation and self-management. Therefore, this could improve patient outcomes and reduce costs. For payers, high-cost specialty drugs can incur financial challenges for the following year. The early projection allows them to manage total drug spending by using various benefit design strategies and ensuring the appropriate use of specialty drugs. There is a lack of a predictive model for general specialty drug use. Our objective was to apply a machine learning-based prediction model to predict specialty drug use among members using claims data.

## MATERIALS AND METHODS

### Study design and data source

We constructed a dataset using administrative claims data from commercially insured individuals in two databases. One of the databases covered members nationally (Commercial Claims and Encounters, or CCAE)* and no members were older than 65 years, while the other covered only a few states and members included those older than 65 years (Regional Data Set). We processed the data of administrative claims from the Regional Data Set in the same manner as the CCAE database. Our study period was from January 1, 2015, to December 31, 2016. 2015 was the Feature Window, and 2016 was the Target Prediction Window. The dataset contained information on demographic characteristics, enrollment, costs, pharmaceutical, and medical utilization. To be qualified for the study cohort, members had to be enrolled in a medical or pharmacy plan for three continuous months immediately before the start of the Target Prediction Window (Figure 1). Members were included in the cohort only if they had no specialty drug related claims under either medical or pharmacy benefits during the Feature Window.

**Figure 1.**
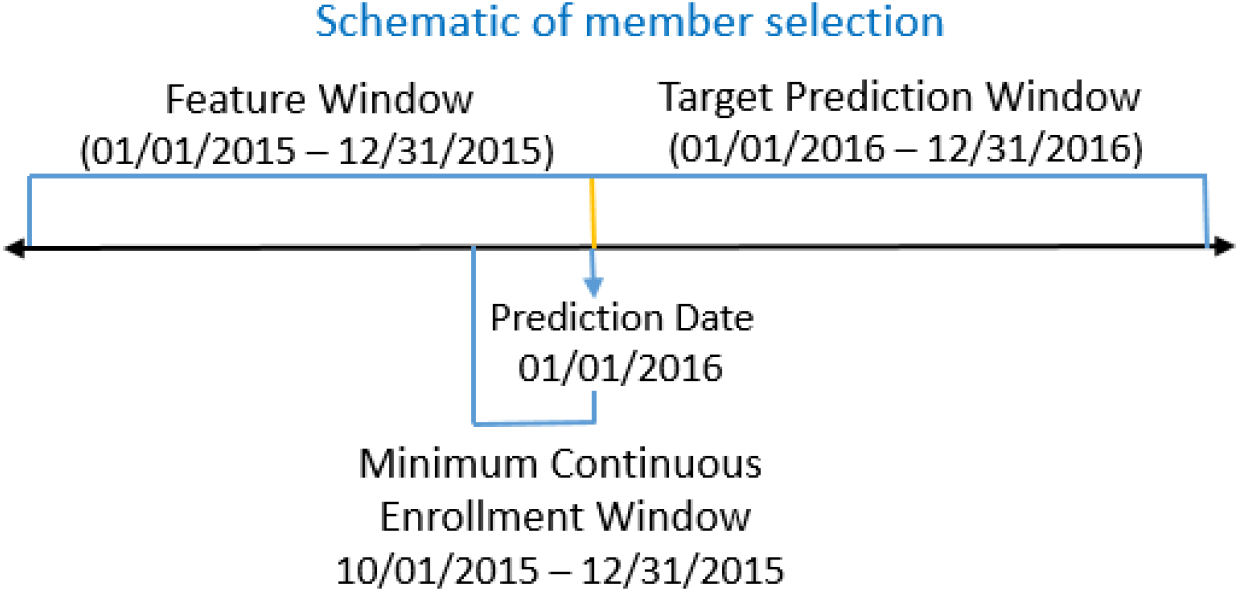
This diagram offers a visual depiction of the model development timeline. The Prediction Date was January 1, 2016. Members who were insured under either a medical or pharmacy plan and who had a minimal continuous enrollment from Oct 1, 2015 through December 31, 2015 were included. All available claims data per patient between January 1, 2015, and December 31, 2015 were used for feature creation.

### Outcome variable: Defining a specialty drug

For the years of 2006-2016, the Centers for Medicare and Medicaid Services (CMS) defined a specialty drug as a product that costs $600 or more for a 30-day supply. The threshold increased to $670 in 2017 [8][9][10]. This is the threshold CMS requires for a product to be put on a specialty tier. We intended to focus our predictions primarily on specialty drug use for treating chronic diseases. Therefore, we further restricted our specialty drug use definition to exclude medications for infertility treatment and erectile dysfunction. The excluded drug codes can be found in Supplementary Table S1.

### Feature candidates

We included 81 features calculated from claims that occurred during the Feature Window of Jan 1, 2015, to December 31, 2015. These 81 features included demographic-related features such as age and gender, disease-related features based on ICD-9-CM [11] or ICD-10-CM diagnosis codes [12], procedure codes [13], NDC and the Anatomical Therapeutic Chemical (ATC) classification codes [14], and member enrollment months. Payment-based features such as monthly payment under the medical or pharmacy plan, member liability amount, copayment, and deductible were assessed and calculated. ICD codes for comorbid conditions included hypertension, chronic heart failure, diabetes, asthma, lipid disorder, depression, arthritis, migraine, gastric acid disorder, chronic renal insufficiency, anemia, end-stage renal disease, chronic hepatitis C, arrhythmia, and osteoporosis (Supplementary Table S2). The patients with cancer, kidney disease, and rheumatoid arthritis (RA) have more co-morbid conditions than population-at-large. They usually have higher rates of hypertension, diabetes, lipid disorder and heart disease. Patients with multiple sclerosis are more likely to have depression and migraines [15]. Utilizations such as outpatient claims, inpatient claims, and emergency department claims were part of features extracted.

### Modeling approach and evaluation

The machine learning algorithm we applied in our dataset was an extreme gradient boosting (XGBoost) classifier [16]. We split our dataset into training (75%, 4.9 million members) and test (25%, 1.6 million members) sets for model training and evaluation. Random grid search and cross validation were used for hyperparameter tuning. Those tuned hyperparameters included maximum depth, minimum child weight, learning rate, gamma, and maximum delta step. The primary metric used to optimize model performance was balanced accuracy. The Youden index was used to choose the optimal threshold for prediction cutoff in order to maximize balanced accuracy [17]. Major Python packages that were used for analyses included Scikit-learn (version 1.15.0) and XGBoost (version 2.0.1). Python version 3.7 was used for data analysis and the model building process.

## RESULTS

### Descriptive analysis results

We performed some preliminary data analysis on the Regional Data Set, which included both members younger and older than 65 years old. We did some analyses on members enrolled in 2015 and having at least three-month continuous enrollment under either a medical or a pharmacy plan. We found that 5.4% of members were using specialty drugs in 2015. In 2016, 7.0% of the same members were taking specialty drugs, which represented a modest increase compared to 2015. We further investigated and found that 62.6% of the members who used specialty drugs in 2015 were still taking specialty drugs in 2016. Meanwhile, among the members who were not taking any specialty drugs, 3.8% became specialty drug users in 2016. We were therefore interested in predicting which enrolled members would become specialty drug users in the next year based on the previous year’s medical and pharmacy history.

Specialty drugs could be covered under either the pharmacy or medical plan. There has been an ongoing discussion of the advantages and disadvantages regarding covering specialty drugs in either of the two plans [18]. Therefore, we compared the specialty drug spending between medical plan and pharmacy plan. First, we examined the total spending on specialty drugs under the pharmacy plan and identified the top 10 most expensive medications based on ATC 4^th^ level in 2016 (Table 1). Then we determined the top 10 most costly ATC 4^th^ level medications under the medical plan in 2016 (Table 2).

**Table 1:**
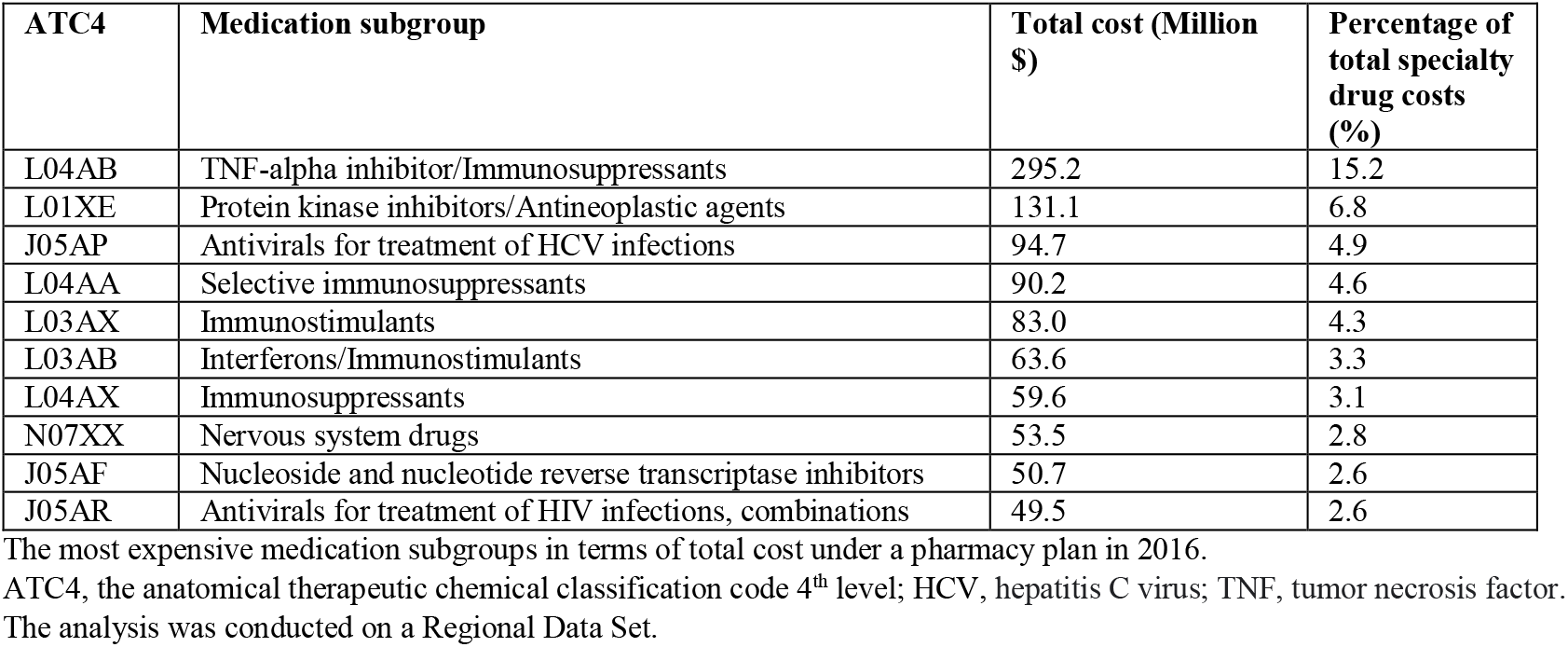
Top 10 most expensive specialty medication subgroups under a pharmacy plan

| ATC4 | Medication subgroup | Total cost (Million \$) | Percentage of total specialty drug costs (%) |
| --- | --- | --- | --- |
| L04AB | TNF-alpha inhibitor/Immunosuppressants | 295.2 | 15.2 |
| L01XE | Protein kinase inhibitors/Antineoplastic agents | 131.1 | 6.8 |
| J05AP | Antivirals for treatment of HCV infections | 94.7 | 4.9 |
| L04AA | Selective immunosuppressants | 90.2 | 4.6 |
| L03AX | Immunostimulants | 83.0 | 4.3 |
| L03AB | Interferons/Immunostimulants | 63.6 | 3.3 |
| L04AX | Immunosuppressants | 59.6 | 3.1 |
| N07XX | Nervous system drugs | 53.5 | 2.8 |
| J05AF | Nucleoside and nucleotide reverse transcriptase inhibitors | 50.7 | 2.6 |
| J05AR | Antivirals for treatment of HIV infections, combinations | 49.5 | 2.6 |

The most expensive medication subgroups in terms of total cost under a pharmacy plan in 2016.
ATC4, the anatomical therapeutic chemical classification code 4<sup>th</sup> level; HCV, hepatitis C virus; TNF, tumor necrosis factor.
The analysis was conducted on a Regional Data Set.

**Table 2:** Top 10 most expensive specialty medication subgroups under a medical plan

| ATC4 | Medication subgroup | Total cost (Million \$) | Percentage of total specialty drug costs (%) |
| --- | --- | --- | --- |
| B02BD | Blood coagulation factors | 198.8 | 14.9 |
| L01XC | Antineoplastic agents | 189.8 | 14.3 |
| L03AA | Colony stimulating factors/Immunostimulants | 63.4 | 4.8 |
| L04AB | TNF-alpha inhibitor | 61.6 | 4.6 |
| L01CD | Taxanes/Antineoplastic agents | 59.4 | 4.5 |
| L04AA | Selective immunosuppressants | 58.4 | 4.4 |
| L04AC | Interleukin inhibitors/Immunosuppressants | 49.8 | 3.7 |
| L01AA | Pyrimidine analogs/Antineoplastic agents | 48.7 | 3.7 |
| L01BC | Nitrogen mustard analogs/Antineoplastic agents | 41.0 | 3.1 |
| L01XX | Antineoplastic agents | 40.2 | 3.0 |

The most expensive medication subgroups in terms of total cost under a medical plan in 2016. ATC4, the anatomical therapeutic chemical classification code 4<sup>th</sup> level; TNF, tumor necrosis factor. The analysis was conducted on a Regional Data Set.

We further investigated the specialty drug spending among various age groups among the members included in the cohort. First, we examined the total cost of specialty drugs under the medical and pharmacy plans. We found a similar pattern under the two plans. There was a substantial increase in specialty drug use after age 40, with the trend being more evident under the pharmacy plan. Specialty drug spending reached a peak for members at around 60 years old (Figure 2A, B&C). Those members in the age group between 50 to 59 years old spent the most under both pharmacy and medical plans in 2016 (Figure 2A, B&C). We also noticed a different pattern between the pharmacy and medical plans. Among members younger than 18, there was considerable more spending on specialty drugs under the pharmacy plan than under the medical plan (Figure 2A, B&C). Since Medicare members represented a small portion of the study cohort, we then investigated the average cost of specialty drugs under the medical and pharmacy plans. The average specialty drug spending was much higher under the medical plan than under the pharmacy plan (Figure 2D, E&F). The average specialty drug spending was highest among members who were between 40 and 60 years old under the medical plan (Figure 2D). However, we observed a slight but constant rise of average specialty drug spending as the age increased under the pharmacy plan (Figure 2E).

**Figure 2.**
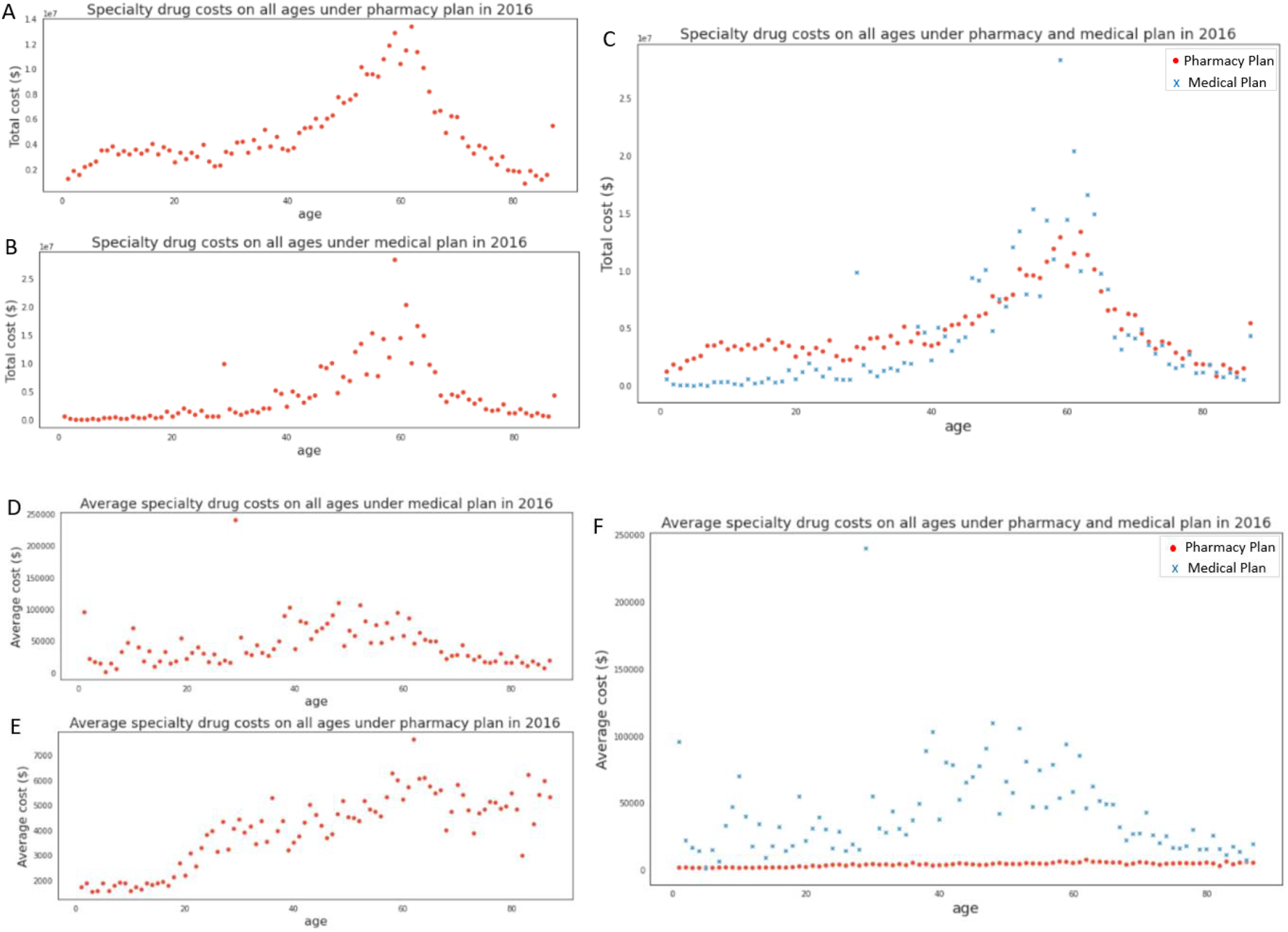
The specialty drug spending pattern among various age groups under pharmacy and medical plans in 2016. (A) The scatter plot of specialty drug spending across the ages under the pharmacy plan. (B) The scatter plot of specialty drug spending across the ages under the medical plan. (C) The combined scatter plot of specialty drug spending across the ages under pharmacy plan and medical plan. This analysis was conducted on the Regional Data Set, covering 1 million members.

A member’s past payment history could be predictive of that member’s future costs, including medication and other medical services. We examined the monthly payment pattern among members covered by either a medical plan or a pharmacy plan. In Figure 3A, we saw that 19% of the members enrolled in a pharmacy plan paid for at least one prescription during each and every month of 2015, which indicated that those members could have some type of chronic diseases and became regular medication users. Seventeen percent of the members paid for a prescription during only one month of 2015. In contrast, among members enrolled in a medical plan, 14% of them paid for at least one covered medication, test, or treatment service during only one month of 2015. Only 4.7% of them paid each and every month in 2015 for any type of services (Figure 3B).

**Figure 3.**
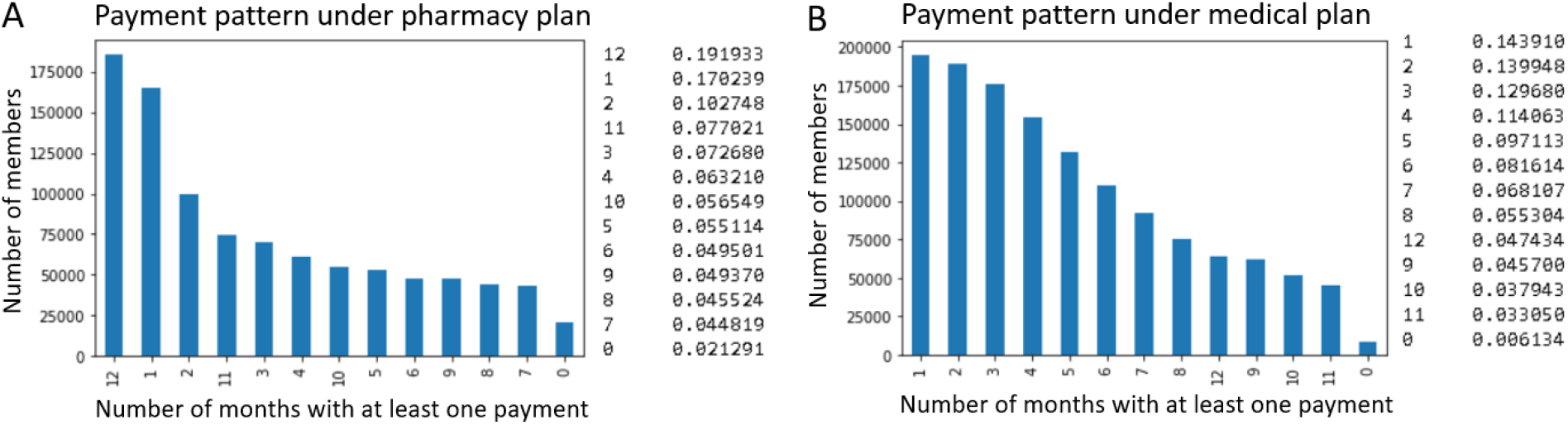
Monthly payment pattern among members covered either medical plan or pharmacy plan in 2015. (A) Number of months with at least one payment by members under the pharmacy plan. (B) Number of months with at least one payment by members under the medical plan. This analysis was conducted on a Regional Data Set. The left column of the legend indicated the number of months with at least one payment. The right column of the legend was percentage of member counts.

### Prediction performance of ML algorithms

The feature importances for the 20 most important features are shown in Figure 4. The most important predictor was age. Monthly payments from the medical and pharmacy plan which contained rich information regarding the pattern of how often and how much patients paid for their medication and medical services were highly predictive (Supplementary Table S2).

**Figure 4.**
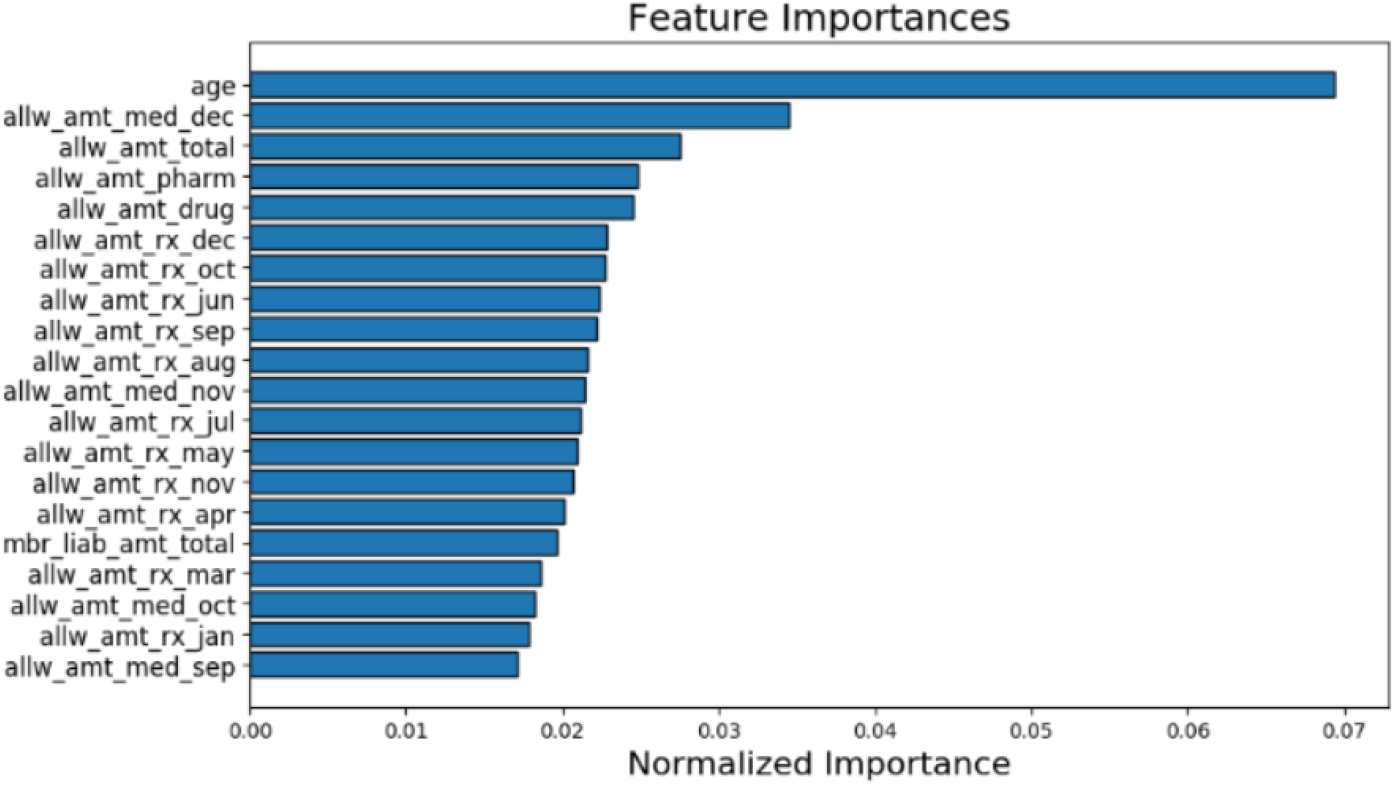
Top 20 most important features for predicting. See Supplemental Table S2 for the feature description of each feature name.

To evaluate model performance, we split the data randomly into train and test sets. We first started with 1 million members in the Regional Data Set, 75 percent of members were in the training set, and 25 percent were in the test set. XGBoost algorithm was fit and tuned to the training data. We then evaluated the model performance on the test set. To improve model generalization, we gradually added members from the CCAE database to get 6.5 million in total. Among the 6.5 million members, most of the member data were from the CCAE database where there were no Medicare patients. Medicare members account for only a small portion of the total members in the study, so we also evaluated the model performance on the test set for those members older than 65. The balanced accuracy for the train set was 73% and that for the test set was 71%, the balanced accuracy for the test set with members older than 65 was 71%.

We then set the best classification threshold based on maximizing balanced accuracy. The balanced accuracy of the model was similar when it was calculated separately on members of all ages and on members older than 65 years (Table 3). The model performance was also similar between the two age groups for the other performance metrics. We present the model’s AUROC curve and precision-recall curve in Figure 5.

**Table 3:** Model performance metrics on test datasets from different age cohorts

| Metrics | Members of all ages | Members with age >=65 |
| --- | --- | --- |
| Prevalence rate | 0.0303 | 0.0363 |
| Balanced accuracy | 0.7107 | 0.7076 |
| Recall | 0.7108 | 0.7077 |
| Specificity | 0.7106 | 0.7075 |
| AUROC | 0.7858 | 0.7930 |
| NPV | 0.9875 | 0.9847 |
| F1 | 0.1294 | 0.1495 |

Numbers were calculated based on threshold cutoffs to optimize specificity and sensitivity on training data. AUROC, area under the receiver operating characteristic curve; NPV, negative predictive value.

**Figure 5.**
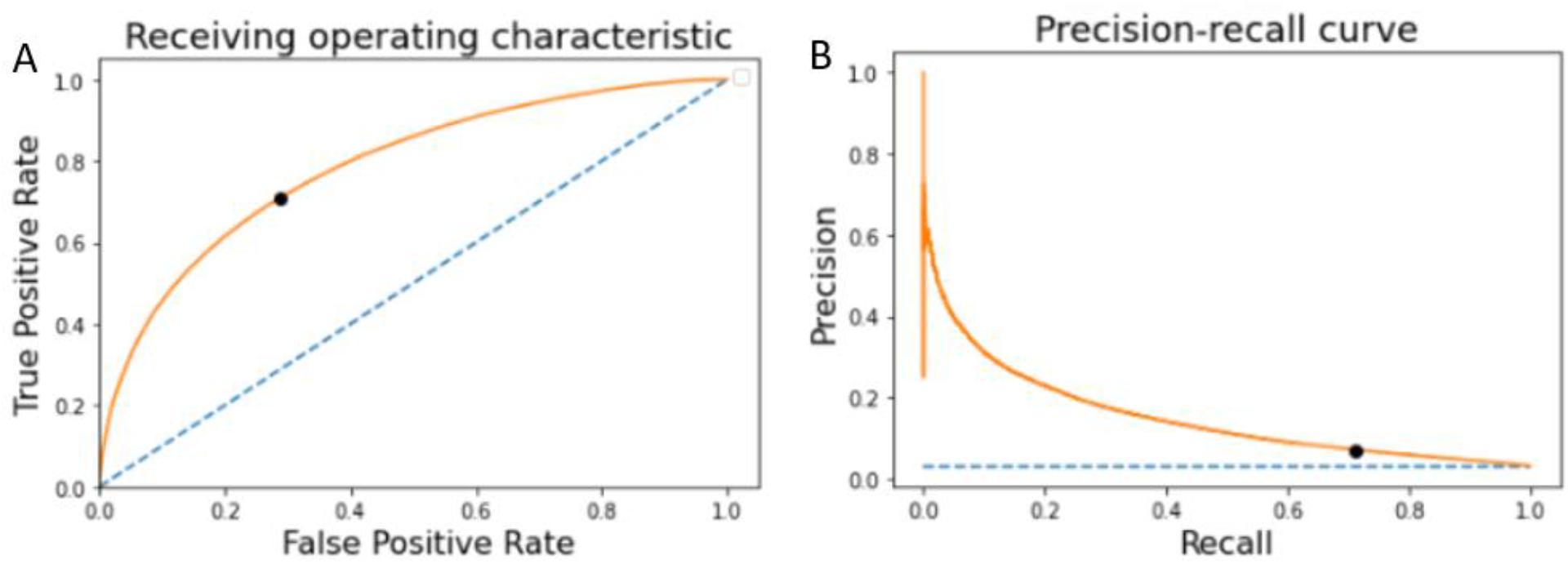
Model performance metrics on the test dataset. (A) Receiving operating characteristic. AUROC = 0.786 (B) Precision-recall curve. AUPRC = 0.151. The blue dashed line indicates baseline random guess performance. The black dot represents the classifier at the chosen, optimal threshold for prediction.

## DISCUSSION

Our predictive model was based on the concept that we can help payers or providers identify the members who may use high cost specialty drugs in the next 12 months. This will allow them to make a forecast on next year’s spending and perform disease management. A care management team can help members access the medication, remind them to take the medication, and help them manage adverse drug events. If members cannot access the specialty drugs, it may cause adverse clinical outcomes and lead to more high cost utilization, such as inpatient or emergency room visits. Proper intervention can help slow down disease progression and improve their quality of life.

This model used both disease- and cost-related features under medical and pharmacy plans to make the prediction. For cost-related features, the monthly payment pattern was highly predictive which was a new discovery in this model. The payment patterns could have a profound indication for members’ disease status and medication use habits. Members who might use specialty drugs usually have some chronic medical conditions, which were also captured in this model. In Table 1 and Table 2, we found that the most expensive specialty drugs in the current study were cancer medication and immunosuppressant. The patients with cancer and RA generally have comorbidities. Four out of ten patients 65 years or older with cancer have at least one other chronic condition. Patients with comorbidity are generally less likely to receive curative treatment for their cancer than those without comorbidity [19]. Due to other coexisting diseases, such as cardiovascular disease (CVD) and infections, RA patients have more increased mortality than the general population [20]. Some studies have shown that patients with RA who are clinical responders to certain anti-rheumatic treatment such as anti-TNF treatment have a lower risk of CVD than nonresponders [21].

There are no similar machine learning models to predict specialty drug use to our knowledge at the time of this writing. Our model achieved comparable performance for members of all ages and those older than 65. In our highly imbalanced classification model (3.0% prevalence rate), the balanced accuracies for all ages and on members older than 65 were both greater than 70%. Our model provides a unique opportunity for providers and payers to identify earlier those members who will become specialty drug users, which may lead to high costs and can be used to treat disease progression or comorbidities. Patients can also benefit from disease management and have improved clinical outcomes. There is a growing financial burden of specialty drug use for payers due to its high-cost characteristic. The early prediction of specialty drug use can help payers manage drug costs and perform benefit design to reduce financial loss.

### Limitations

In our study, we applied CMS’ definition for specialty drugs to our data set. Each commercial health plan has its own specialty drug tier definition, which may or may not overlap with the CMS definition. There were no social factors-related features included in this model. Income, race, environment could impact the overall health in a certain population, and some drugs are more or less responsive to some members due to genetic variations. Many specialty drug users have complex disease status and comorbidities that make their drug regime very complicated. Whether or not a doctor will prescribe a specialty drug was based on many factors, such as disease progression, current medication taken by the patient, financial status, patient’s health condition, and clinician’s knowledge. With clinical data, we might extract better predictors to make the model more adaptable to clinical settings.

### Next steps

We plan to build a specialty drug regression model to predict the total costs of specialty drug use under pharmacy plan or medical plan at each member level. With the prediction of total costs in either the pharmacy or medical plan, we can have a better picture of increased cost in each plan, allowing payers or providers to evaluate and develop targeted intervention strategies and create a framework to administer outcomes-based contracts.

## CONCLUSIONS

The cost of prescription drugs is a leading health policy issue in the United States. The spending on specialty drugs is on the rise every year. The ability to predict the members who might use specialty drugs in the next 12 months meets benefit managers’ interest in monitoring and containing their utilization. Care management can focus on making sure that members who most benefit from taking specialty drugs can access them. The proper use of specialty drugs may help avoid more expensive procedures in the future.

## Data Availability

The data used for this manuscript are commercially licensed from the CCAE database and not publically
available. The independent dataset contains sensitive personal health information and is therefore also
not publically available.

## ACKNOWLEDGMENTS

The authors thank Natalie Benner, Mackenzie DeBoer, Jessica Sears for helpful discussion.

## DATA AVAILABILITY

The data used for this manuscript are commercially licensed from the CCAE database and not publically available. The Regional Data Set contains de-identified health information and is not publically available due to ownership restrictions.

## SUPPLEMENTARY MATERIAL

**Supplementary Table S1:** Infertility and ED codes

| Disease | Diagnostic code | Procedure code | ATC4 code | NDC |
| --- | --- | --- | --- | --- |
| Infertility | (N97\|628\|N46\|606\|E291\|2572\|E288\|2568\|E2839\|25639).* | S0122\|S0126\|S0128\|S0132\|J0725\|J3355\|J3490 | N/A | (555668505\|440871225\|498840701\|000236150\|000236151\|550560806\|550560808\|550560818\|547714311\|000520313\|000520316\|000520326\|440871115\|440871116\|440871117\|440871118\|555667501\|555661502\|555661501\|555661700\|440871150\|440870250\|555667185\|555667125\|000520315\|548684997\|695430373\|548684230\|548684250\|440878090\|440877075\|440877150\|003003612).* |
| Erectile dysfunction | (N52\|60784).* | N/A | G04BE | N/A |

\* represents zero or more characters

**Supplementary Table S2:**
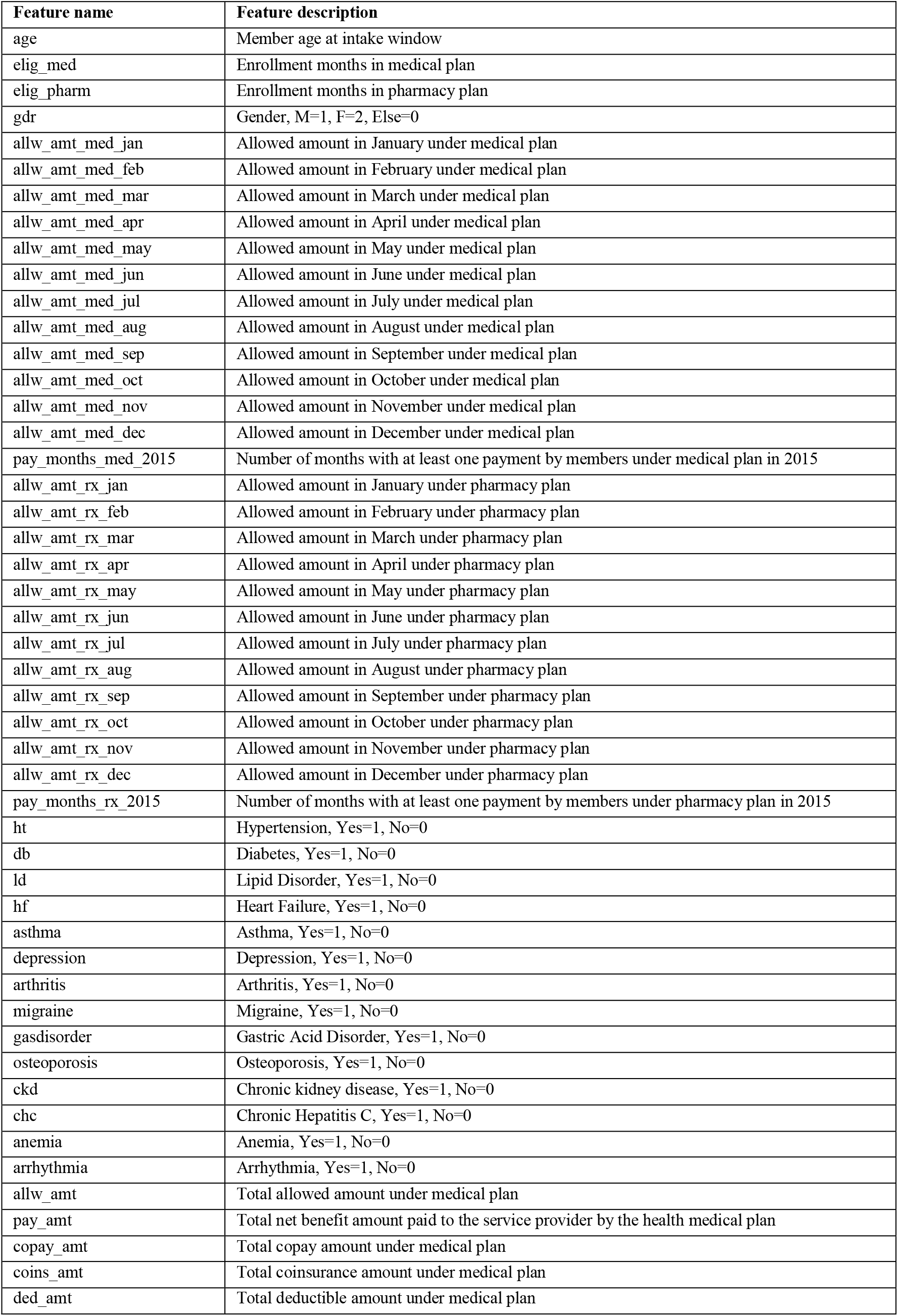
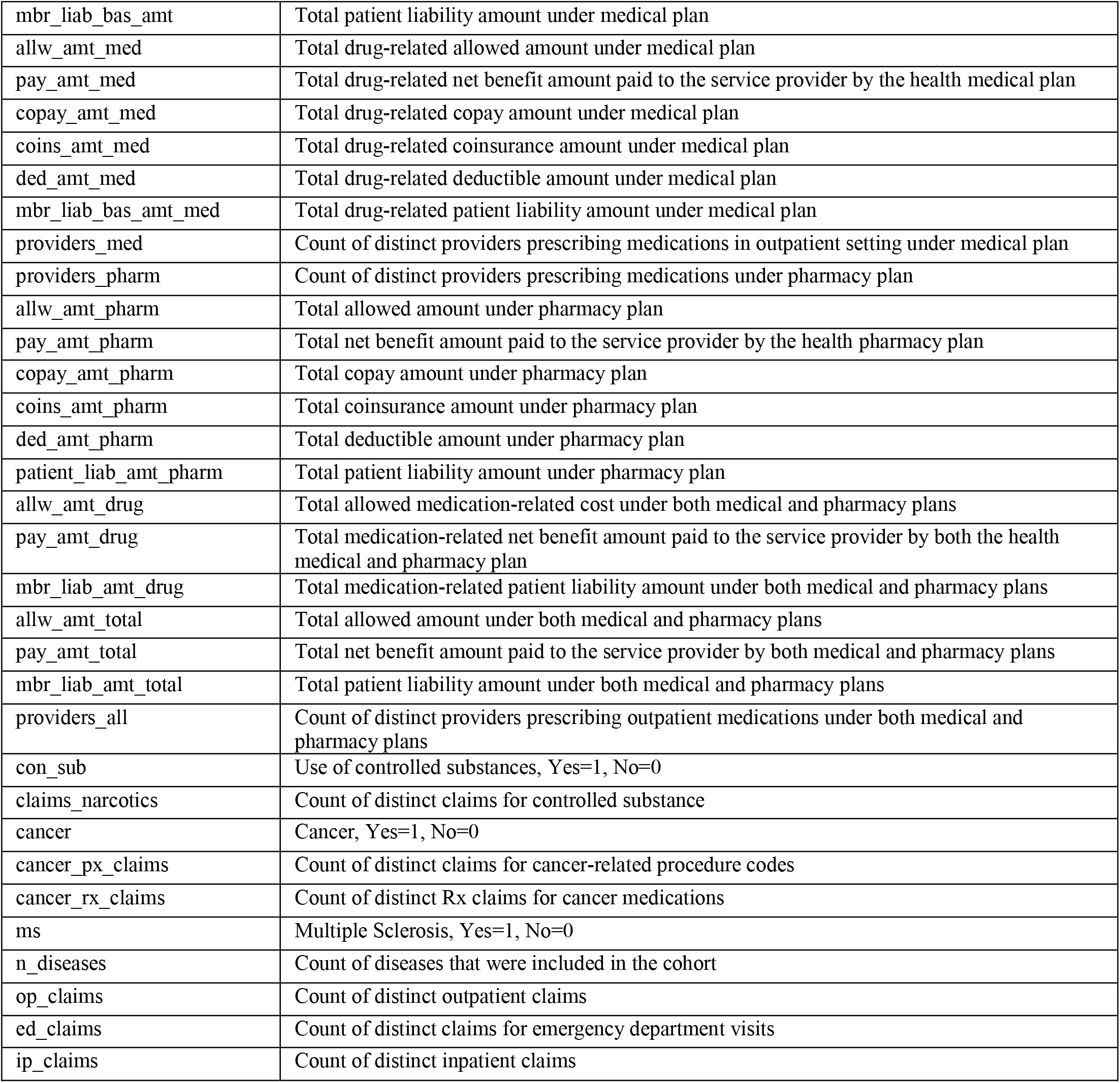
Feature names and definitions (n=81)

| Feature name | Feature description |
| --- | --- |
| age | Member age at intake window |
| elig_med | Enrollment months in medical plan |
| elig_pharm | Enrollment months in pharmacy plan |
| gdr | Gender, M=1, F=2, Else=0 |
| allw_amt_med_jan | Allowed amount in January under medical plan |
| allw_amt_med_feb | Allowed amount in February under medical plan |
| allw_amt_med_mar | Allowed amount in March under medical plan |
| allw_amt_med_apr | Allowed amount in April under medical plan |
| allw_amt_med_may | Allowed amount in May under medical plan |
| allw_amt_med_jun | Allowed amount in June under medical plan |
| allw_amt_med_jul | Allowed amount in July under medical plan |
| allw_amt_med_aug | Allowed amount in August under medical plan |
| allw_amt_med_sep | Allowed amount in September under medical plan |
| allw_amt_med_oct | Allowed amount in October under medical plan |
| allw_amt_med_nov | Allowed amount in November under medical plan |
| allw_amt_med_dec | Allowed amount in December under medical plan |
| pay_months_med_2015 | Number of months with at least one payment by members under medical plan in 2015 |
| allw_amt_rx_jan | Allowed amount in January under pharmacy plan |
| allw_amt_rx_feb | Allowed amount in February under pharmacy plan |
| allw_amt_rx_mar | Allowed amount in March under pharmacy plan |
| allw_amt_rx_apr | Allowed amount in April under pharmacy plan |
| allw_amt_rx_may | Allowed amount in May under pharmacy plan |
| allw_amt_rx_jun | Allowed amount in June under pharmacy plan |
| allw_amt_rx_jul | Allowed amount in July under pharmacy plan |
| allw_amt_rx_aug | Allowed amount in August under pharmacy plan |
| allw_amt_rx_sep | Allowed amount in September under pharmacy plan |
| allw_amt_rx_oct | Allowed amount in October under pharmacy plan |
| allw_amt_rx_nov | Allowed amount in November under pharmacy plan |
| allw_amt_rx_dec | Allowed amount in December under pharmacy plan |
| pay_months_rx_2015 | Number of months with at least one payment by members under pharmacy plan in 2015 |
| ht | Hypertension, Yes=1, No=0 |
| db | Diabetes, Yes=1, No=0 |
| ld | Lipid Disorder, Yes=1, No=0 |
| hf | Heart Failure, Yes=1, No=0 |
| asthma | Asthma, Yes=1, No=0 |
| depression | Depression, Yes=1, No=0 |
| arthritis | Arthritis, Yes=1, No=0 |
| migraine | Migraine, Yes=1, No=0 |
| gasdisorder | Gastric Acid Disorder, Yes=1, No=0 |
| osteoporosis | Osteoporosis, Yes=1, No=0 |
| ckd | Chronic kidney disease, Yes=1, No=0 |
| chc | Chronic Hepatitis C, Yes=1, No=0 |
| anemia | Anemia, Yes=1, No=0 |
| arrhythmia | Arrhythmia, Yes=1, No=0 |
| allw_amt | Total allowed amount under medical plan |
| pay_amt | Total net benefit amount paid to the service provider by the health medical plan |
| copay_amt | Total copay amount under medical plan |
| coins_amt | Total coinsurance amount under medical plan |
| ded_amt | Total deductible amount under medical plan |
| mbr_liab_bas_amt | Total patient liability amount under medical plan |
| allw_amt_med | Total drug-related allowed amount under medical plan |
| pay_amt_med | Total drug-related net benefit amount paid to the service provider by the health medical plan |
| copay_amt_med | Total drug-related copay amount under medical plan |
| coins_amt_med | Total drug-related coinsurance amount under medical plan |
| ded_amt_med | Total drug-related deductible amount under medical plan |
| mbr_liab_bas_amt_med | Total drug-related patient liability amount under medical plan |
| providers_med | Count of distinct providers prescribing medications in outpatient setting under medical plan |
| providers_pharm | Count of distinct providers prescribing medications under pharmacy plan |
| allw_amt_pharm | Total allowed amount under pharmacy plan |
| pay_amt_pharm | Total net benefit amount paid to the service provider by the health pharmacy plan |
| copay_amt_pharm | Total copay amount under pharmacy plan |
| coins_amt_pharm | Total coinsurance amount under pharmacy plan |
| ded_amt_pharm | Total deductible amount under pharmacy plan |
| patient_liab_amt_pharm | Total patient liability amount under pharmacy plan |
| allw_amt_drug | Total allowed medication-related cost under both medical and pharmacy plans |
| pay_amt_drug | Total medication-related net benefit amount paid to the service provider by both the health medical and pharmacy plan |
| mbr_liab_amt_drug | Total medication-related patient liability amount under both medical and pharmacy plans |
| allw_amt_total | Total allowed amount under both medical and pharmacy plans |
| pay_amt_total | Total net benefit amount paid to the service provider by both medical and pharmacy plans |
| mbr_liab_amt_total | Total patient liability amount under both medical and pharmacy plans |
| providers_all | Count of distinct providers prescribing outpatient medications under both medical and pharmacy plans |
| con_sub | Use of controlled substances, Yes=1, No=0 |
| claims_narcotics | Count of distinct claims for controlled substance |
| cancer | Cancer, Yes=1, No=0 |
| cancer_px_claims | Count of distinct claims for cancer-related procedure codes |
| cancer_rx_claims | Count of distinct Rx claims for cancer medications |
| ms | Multiple Sclerosis, Yes=1, No=0 |
| n_diseases | Count of diseases that were included in the cohort |
| op_claims | Count of distinct outpatient claims |
| ed_claims | Count of distinct claims for emergency department visits |
| ip_claims | Count of distinct inpatient claims |

